# A Single-Tube Colorimetric Loop-Mediated Isothermal Amplification for Rapid Detection of SARS-CoV-2 RNA

**DOI:** 10.1101/2023.08.10.23293869

**Authors:** Sayamon Hongjaisee, Nang Kham-Kjing, Piyagorn Musikul, Wannaporn Daengkaokhew, Nuntita Kongson, Ratchadakorn Guntala, Nitipoom Jaiyapan, Enos Kline, Nuttada Panpradist, Nicole Ngo-Giang-Huong, Woottichai Khamduang

**Affiliations:** Research Institute for Health Sciences, Chiang Mai University, Chiang Mai, Thailand; LUCENT International Collaboration, Faculty of Associated Medical Sciences, Chiang Mai University, Chiang Mai, Thailand; Department of Medical Technology, Faculty of Associated Medical Sciences, Chiang Mai University, Chiang Mai, Thailand; School of Medical Sciences, University of Phayao, Phayao, Thailand; Department of Bioengineering, University of Washington, Seattle, Washington, United States; Department of Global Health, University of Washington, Seattle, Washington, United States; Maladies Infectieuses et Vecteurs: Écologie, Génétique, Évolution et Contrôle (MIVEGEC), Agropolis Uni-versity Montpellier, Centre National de la Recherche Scientifique (CNRS), Institut de Recherche Pour le Développement (IRD), Montpellier, France; International Joint Laboratory PRESTO, Chiang Mai, Thailand

**Keywords:** COVID-19, SARS-CoV-2, RT-LAMP, Loop-mediated isothermal amplification, Hydroxynaphthol blue, Cresol red

## Abstract

Since SARS-CoV-2 is a highly transmissible virus, a rapid and accurate diagnostic method is necessary to prevent virus spread. We aimed to develop and evaluate a new rapid colorimetric reverse transcription loop-mediated isothermal amplification (RT-LAMP) assay for SARS-CoV-2 detection in a single closed-tube. Nasopharyngeal and throat swabs collected from at-risk individuals testing for SARS-CoV-2 were used to assess the sensitivity and specificity of a new RT-LAMP assay against a commercial qRT-PCR assay. Total RNA extracts were submitted to the RT-LAMP reaction under optimal conditions and amplified at 65°C for 30 minutes using three sets of specific primers targeting the nucleocapsid gene. The reaction was detected using two different indicator dyes, hydroxynaphthol blue (HNB) and cresol red. A total of 82 samples were used for detection with HNB and 94 samples with cresol red, and results were compared with the qRT-PCR assay. The sensitivity of the RT-LAMP-based HNB assay was 92.1% and the specificity was 93.2%. The sensitivity of the RT-LAMP-based cresol red assay was 80.3%, and the specificity was 97%. This colorimetric feature makes this assay highly accessible, low-cost, and user-friendly, which can be deployed for massive scale-up and rapid diagnosis of SARS-CoV-2 infection, particularly in low-resource settings.

## 1. Introduction

The outbreak of a novel severe acute respiratory syndrome coronavirus (SARS-CoV-2) in late 2019 has led to a global COVID-19 pandemic. As of May 8, 2023, the number of global coronavirus cases had exceeded 765 million, resulting in 6.9 million fatalities across nearly 200 countries [1]. The COVID-19 pandemic has emphasized the need for rapid and accurate diagnostic methods for the detection of SARS-CoV-2 to limit the spread of the virus. Nowadays, several rapid antigen test kits (ATKs) can be used for massive detection and are cheap compared to molecular tests. However, ATK has a lower sensitivity than molecular tests [2]. Real-time quantitative reverse transcription polymerase chain reaction (qRT-PCR) has been widely adopted as the gold standard for the molecular diagnosis of SARS-CoV-2 due to its high specificity and sensitivity. However, most real-time RT-PCR assays provide results in over an hour. Furthermore, they require expensive equipment and specialized laboratory facilities, which hinder their implementation, particularly in remote and resource-limited settings.

One recent technique, reverse transcription loop-mediated isothermal amplification (RT-LAMP), has emerged as a promising technique for the rapid detection of various pathogens, including influenza [3], Middle East respiratory syndrome coronavirus [4], hepatitis C virus [5], and Ebola [6]. Unlike PCR, LAMP operates under isothermal conditions and can be performed using a regular water bath or heat block that maintains a constant temperature. The LAMP reaction can be completed within a short time frame (15–60 minutes) and offers high specificity [7]. Several RT-LAMP-based assays have also been designed for SARS-CoV-2 detection, targeting different regions of the viral genome with distinct primer sets [8-13]. Most of these assays utilized relatively expensive commercial reagents for colorimetric RT-LAMP reactions [8,10,12]. Some studies have employed turbidity or fluorescence detection, which still requires specialized and costly equipment not available in resource-limited settings [9,11,12]. Other less costly detection methods have been proposed to remove the need for expensive equipment or instruments [14,15].

We present herein the development of a rapid colorimetric RT-LAMP assay for the detection of SARS-CoV-2 in a single closed-tube and the evaluation of its sensitivity and specificity on clinical samples. A key advantage of our assay is that the reaction color change can be easily observed with the naked eye, eliminating the need for specialized equipment or complex detection methods.

## 2. Materials and Methods

### 2.1. Sample Collection

Nasopharyngeal and throat swabs were collected from 176 symptomatic individuals who underwent SARS-CoV-2 testing at the Faculty of Associated Medical Sciences-Clinical Service Center (AMS-CSC), Chiang Mai University (CMU), between May 2020 and March 2022. All samples used in this study were residual samples. This study was approved by the Faculty of Associated Medical Sciences Ethic Committee (AMSEC-64EM-028) and authorized by the Institutional Biosafety Committee of the Research Institute for Health Sciences, CMU (CMUIBC0363002).

### 2.2. RNA extraction

Total RNA was extracted from 200 μL of swab samples using either the QIAamp Viral RNA Mini Kit (QIAGEN, Germany) or the Nucleic Acid Extraction Kit (Zybio, China), following the manufacturer’s instructions. From the resulting 60 μL total RNA extract, 5μL were subjected to the qRT-PCR SARS-CoV-2 assay (DaAn GENE Co., Ltd., China) on the automated abCyclerQ instrument (ATI Biotech, Singapore), where the cycle threshold (CT) value above 40 is considered undetectable. Another 5 μL of viral RNA was tested for SARS-CoV-2 RNA with the RT-LAMP assay.

### 2.3. RT-LAMP Assay

The RT-LAMP primers targeting the SARS-CoV-2 nucleocapsid gene (N gene) were derived from the previous study [16]. Two outer primers (F3 and B3), two inner primers (FIP and BIP), and two loop primers (LF and LB) were used in the RT-LAMP reaction. The oligonucleotides were synthesized by Integrated DNA Technologies (Coralville, IA, USA).

#### 2.3.1. RT-LAMP assay based on HNB

Colorimetric detection with a metal indicator dye, hydroxynaphthol blue (HNB): a 25 μL reaction was prepared as follows: 1X isothermal amplification buffer (New England Biolabs, MA, USA), 8 mM MgSO_4_, 1.4 mM deoxynucleotide (dNTP) mix, 1.6 mM of FIP and BIP primers, 0.2 mM F3 and B3 primers, 0.4 mM LF and LB primers, 7.5 U WarmStart RTx Reverse Transcriptase (New England Biolabs, MA, USA), 8 U *Bst* 2.0 WarmStart DNA Polymerase (New England Biolabs, MA, USA), 120 μM hydroxynaphthol blue-HNB (Honeywell, Charlotte, North Carolina, USA), and 5 μL RNA template. The amplification reaction was carried out at 65°C for 30 minutes and then terminated at 80°C for 10 minutes. Positive results were indicated by a color change to sky blue, while negative results retained the purple color. To confirm the results, 5 μL of RT-LAMP products were electrophoresed on a 2% w/v agarose gel, stained with RedSafe (iNtRON Biotechnology, Gyeonggi-do, Korea), and bands were visualized with a UV transilluminator. A positive RT-LAMP product displayed a characteristic ladder-like pattern with multiple bands of varying sizes. HNB is a metal ion indicator commonly used in the RT-LAMP reaction. It undergoes a color change from purple to sky blue in the presence of magnesium pyrophosphate, a byproduct of the RT-LAMP reaction [17]. A potential limitation of HNB is that the color change from purple to sky blue may be challenging to discriminate with the naked eye, particularly in certain lighting conditions or for individuals with color vision deficiencies. To address this issue, we used cresol red dye as an alternative indicator in the RT-LAMP assay.

#### 2.3.2. RT-LAMP assay based on cresol red

Colorimetric detection with a pH indicator dye, cresol red: A 25 μL reaction was prepared as follows: 10 mM (NH4)_2_SO_4_, 50 mM KCl, 0.1% v/v Tween-20, 8 mM MgSO_4_, 1.4 mM dNTP mix. The pH of the mixture was adjusted to 8–9 using 1 M KOH, as determined by pH paper. Primers and enzymes were added to the reaction at the same concentrations as the HNB-based assay. In this reaction, 100 μM cresol red was included instead of HNB as the pH indicator dye. The amplification reaction was performed under the same conditions as the HNB-based assay. Cresol red undergoes a color transition from pink to yellow due to a decrease in pH level following the release of protons during amplification [15]. A color change to yellow indicated a positive result, while a pink color indicated a negative result.

## 3. Results

### 3.1. Optimization of RT-LAMP assay based on HNB

The RT-LAMP assay based on HNB dye was optimized by investigating different parameters, including amplification time, MgSO_4_ and HNB dye concentrations. Figure 1 depicts the results of these optimization experiments. To determine the optimal amplification time, the RT-LAMP assay was conducted at 10-minute intervals, from 20 to 60 minutes at 65°C. In the positive control (Figure 1A), a color change from purple to sky blue was observed from 20 minutes onward. However, non-specific amplifications resulting in a color change were observed in the no template control (NTC) after 30 minutes, as confirmed through gel electrophoresis (data not shown). Therefore, a 30-minute amplification time was deemed optimal since it provided the longest reaction time without any color change observed in the NTC.

**Figure 1.**
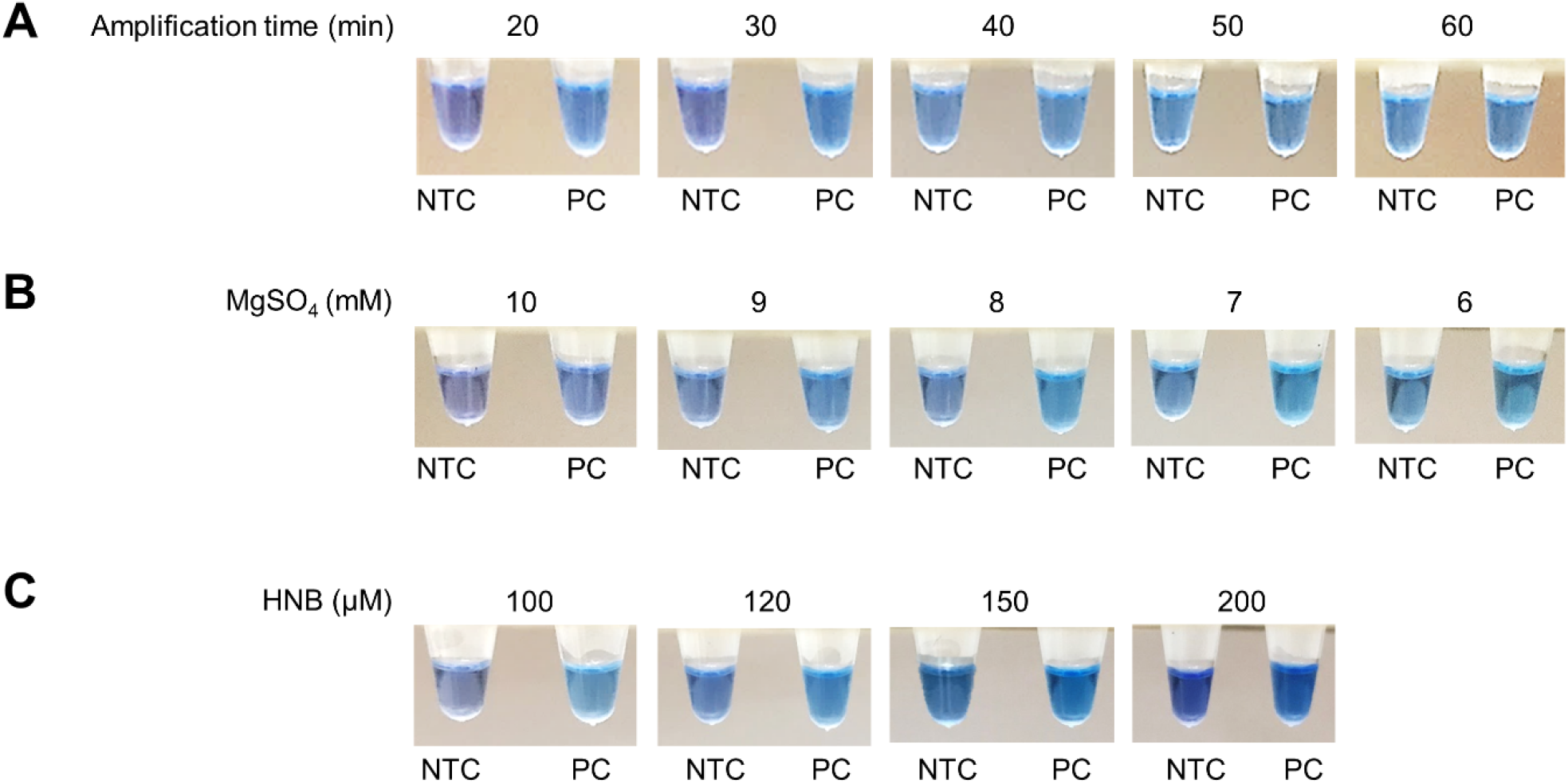
Optimization of RT-LAMP assay based on HNB by varying the amplification time (A), MgSO_4_ concentrations (B), and HNB concentrations (C), using no template control (NTC) and positive control (PC).

Subsequently, the concentrations of MgSO_4_ (ranging from 6–10 mM) and HNB dye (100, 120, 150, and 200 μM) were assessed at 65°C for 30 minutes, followed by inactivation at 80°C for 10 minutes. It was observed that a distinct color change from purple (indicating a negative result) to sky blue (indicating a positive result) occurred at 8 mM MgSO_4_ (Figure 1B) and 120 μM HNB dye (Figure 1C). Therefore, the optimal conditions for the RT-LAMP assay based on HNB were determined as 8 mM MgSO_4_ and 120 μM HNB with an amplification time of 30 minutes at a temperature of 65°C.

### 3.2. Evaluation of the developed colorimetric RT-LAMP assay based on HNB in clinical samples

To evaluate the developed colorimetric SARS-CoV-2 RT-LAMP assay using HNB, a total of 82 clinical samples collected between May 2020 and December 2021 were used for the detection of SARS-CoV-2 RNA. Representative positive and negative results are shown in figure 2. Of these samples, 38 samples were tested positive and 44 samples tested negative by qRT-PCR. Of the 38 positive samples, 35 were tested positive with the colorimetric RT-LAMP based on HNB (Table 1). Three samples that showed false-negative results had CT values of 29.36, 34.35, and 36.32, indicating low SARS-CoV-2 RNA levels in these samples. In this series of clinical samples, the RT-LAMP based on HNB showed a sensitivity for the detection of SARS-CoV-2 of 92.1% and a specificity of 93.2% as compared to the qRT-PCR assay.

**Table 1.** Evaluation of colorimetric RT-LAMP assays on clinical specimens.

| Assays |  | RT-LAMP |  |  |  |
| --- | --- | --- | --- | --- | --- |
|  |  | RT-LAMP<br>based on HNB (N=82) |  | RT-LAMP<br>based on cresol red (N=94) |  |
|  |  | Positive | Negative | Positive | Negative |
| qRT-PCR | Positive | 35<br>(92.1%) | 3<br>(7.9%) | 49<br>(80.3%) | 12<br>(19.7%) |
|  | Negative | 3<br>(6.8%) | 41<br>(93.2%) | 1<br>(3.0%) | 32<br>(97.0%) |

**Figure 2.**
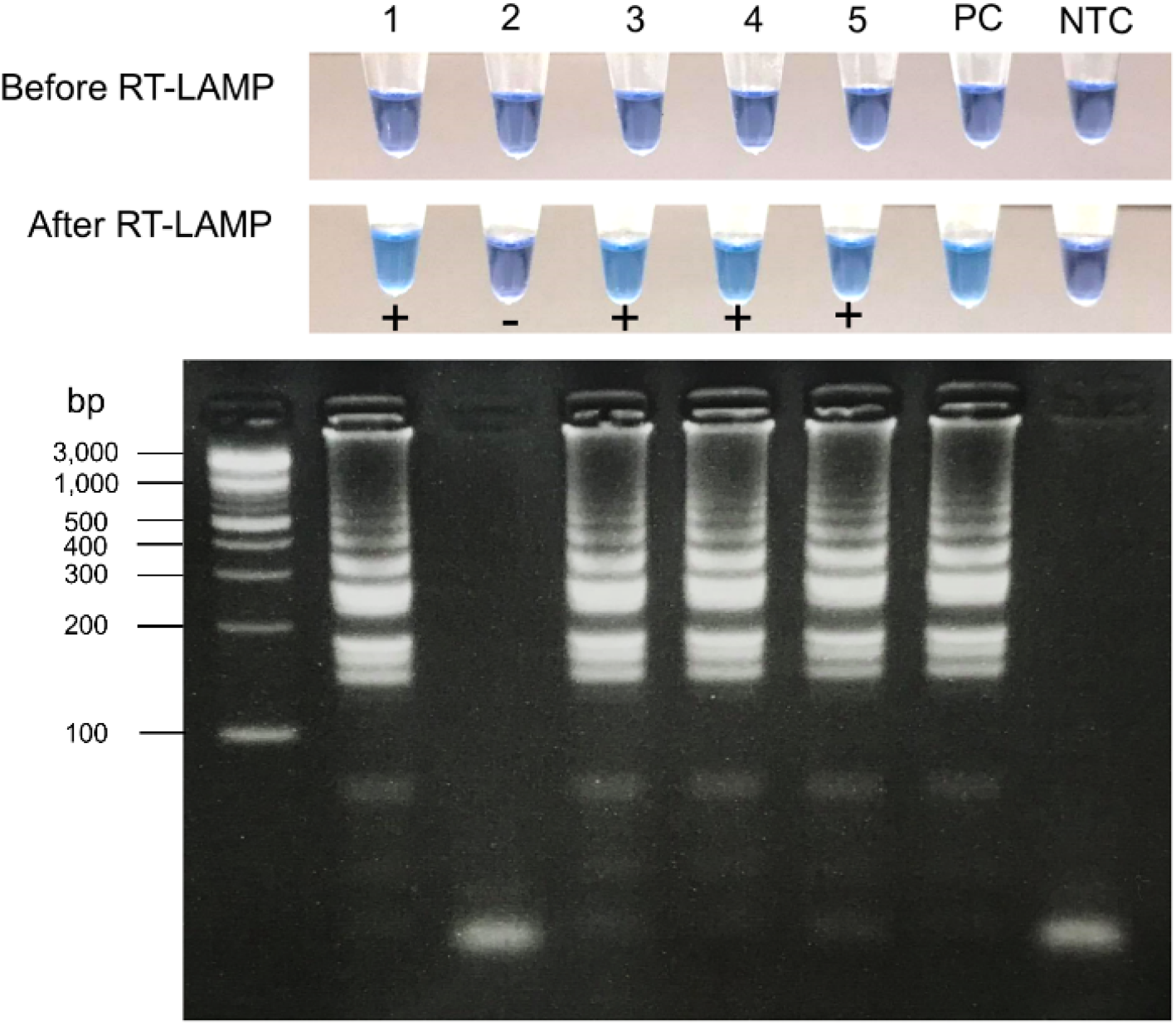
SARS-CoV-2 detection in clinical samples by colorimetric RT-LAMP assay based on HNB. A color change from purple to sky blue was only observed in tubes containing SARS-CoV-2 RNA (no. 1, 3, 4, and 5), whereas the negative sample remained purple (no. 2). The RT-LAMP products have the same gel electrophoresis pattern and correlate with the changes in color. The gel electrophoresis results revealed ladder-like band patterns in SARS-CoV-2 samples but not in negative samples. PC, positive control; NTC, no template control.

### 3.3. Evaluation of the developed colorimetric RT-LAMP assay based on cresol red in clinical samples

The sensitivity and specificity of the colorimetric RT-LAMP based on cresol red for the detection of SARS-CoV-2 RNA were assessed with 94 clinical samples collected between June 2021 and March 2022. Of the 61 samples tested positive by qRT-PCR, 49 were positive by RT-LAMP based on cresol red, and 12 were negative. Among the 12 false-negative samples, 8 samples had a CT value higher than 35 by the qRT-PCR assay, indicating very low SARS-CoV-2 RNA levels, and 4 samples had a CT value below 35 (*i.e*., 18.25, 19.84, 21.25 and 21.58). This assay showed a sensitivity of 80.3% and a specificity of 97.0% (Table 1). Gel electrophoresis was also used to confirm the results (figure 3).

**Figure 3.**
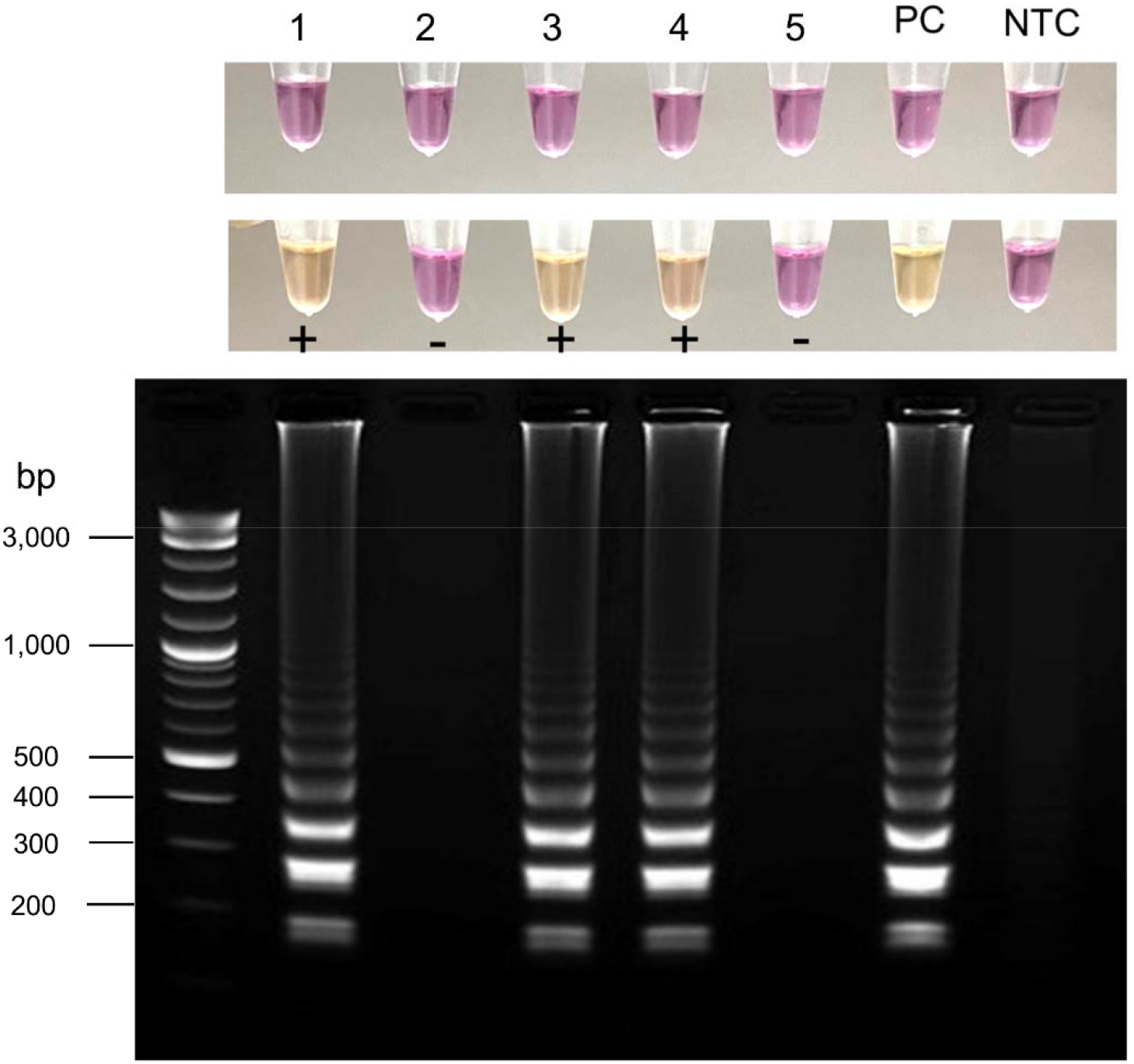
SARS-CoV-2 detection in clinical samples using RT-LAMP with cresol red-based colorimetric detection. A color change from pink to yellow was observed in tubes containing SARS-CoV-2 RNA (no. 1, 3, and 4), while negative samples remained pink (no. 2, and 5). The electrophoresis pattern of the RT-LAMP products is in the same order as the tubes. The results showed the presence of typical ladder-like band patterns in SARS-CoV-2 samples and no ladder-like pattern in negative samples. PC, positive control; NTC, no template control.

## 4. Discussion

We describe here a RT-LAMP assay for SARS-CoV-2 that can be used with two different colorimetric detections: a metal indicator dye (HNB) and a pH indicator dye (cresol red). This assay offers several advantages over qRT-PCR or other RT-LAMP assays using other reporting platforms such as fluorescence dyes [18-20], commercialized colorimetric LAMP [14], turbidity measurement [11], and lateral flow strips [21,22].

First, our assay is simple to perform. The reaction is carried out in a single, closed tube placed in a simple temperature-controlled device and read with the naked eye, making it suitable for resource-limited settings where access to sophisticated or specialized instruments is limited. Second, the results are obtained within 40 minutes, which is crucial for making timely decisions and effective management of infected individuals. Third, the cost of the assay is below 15 USD. Furthermore, this assay has acceptable sensitivity and specificity to detect SARS-CoV-2 RNA.

Detection with HNB is favored due to its simplicity, cost-effectiveness, and safety [23]. However, the color transition of HNB may be difficult to distinguish with the naked eye. The color change in cresol red is more easily distinguishable, as it requires only a minimal change in pH level to trigger a color change from pink to yellow [15]. Consequently, the results can be interpreted more readily and confidently with the cresol red indicator. Detection with HNB gave a sensitivity of 92% and a specificity of 93%. Three false-negative results may be due to the high CT value, indicating a low viral load. Detection with cresol red gave a lower sensitivity of 80.3% and high specificity of 97%. This may be due to the low amount of virus (8 of 12 samples with false-negative results had a CT higher than 35) and/or the different variants circulating during the period of sample collection from June 2021 through March 2022, which may not have been detected. Indeed, mutations in the N gene region corresponding to primer binding sites can affect the detection of SARS-CoV-2 RNA [24,25]. However, we cannot verify or refute this hypothesis since the sequencing of these samples could not be done in this study. Furthermore, four samples exhibited false-positive reactions in both RT-LAMP assays, which could potentially be attributed to cross-contamination. The use of multiple primers in the RT-LAMP assay can lead to the production of a large amount of LAMP products, thereby increasing the risk of cross-contamination and false-positive reactions [26]. Overall, the RT-LAMP assay developed in this study revealed sensitivity and specificity comparable to what was reported by Dao Thi VL et al. using different primer sets to evaluate clinical samples [27]. Ultimately, the choice between HNB and cresol red as indicators in the RT-LAMP assay will depend on user preferences, specific requirements, and resource availability.

This study has some limitations. Firstly, the number of clinical samples tested was relatively low, which may affect the generalizability of the assay’s performance. Secondly, internal control, which could have provided better quality control and detection of potential inhibitory factors, was not included in the assay. Thirdly, the primers used in the study targeted only the N gene, which may limit the assay’s ability to detect viruses with mutations in the primer regions. However, the N gene is highly conserved and abundant. Lastly, the specificity of the developed assay against other related pathogens was not evaluated. To address these limitations and improve the assay’s performance, further studies with larger sample sizes will help establish the robustness and reliability of the assay. Other possible improvements include the addition of an internal control, carrying out a separate reaction with specific primers for RNaseP, targeting multiple gene regions to avoid the risk of false-negatives if mutations occur in one particular region, and increasing the sensitivity and specificity of the assay.

## 5. Conclusions

The RT-LAMP assay using HNB and cresol red dye as visual indicators for SARS-CoV-2 detection represents a great promise for rapid and accurate diagnosis. This assay combines the advantages of simplicity, sensitivity, and rapidity, enabling the detection of SARS-CoV-2 in a cost-effective and timely manner. This assay also provides significant potential for addressing the demand for rapid diagnostics and controlling the spread of SARS-CoV-2.

## Author Contributions

Conceptualization, W.K., S.H., E.K. and N.P.; methodology, N.K.K., P.M., W.D., N.K., R.G. and N.J.; validation, W.K. and S.H.; formal analysis, S.H., N.K.K., P.M., W.D., N.K., R.G. and N.J. ; investigation, W.K., S.H. and N.N.; resources, W.K., S.H. and N.N.; data curation, S.H., N.K.K., P.M., W.D., N.K., R.G. and N.J.; writing—original draft preparation, S.H., N.K.K., W.K. and N.N.; writing—review and editing, S.H., N.K.K., P.M., W.D., N.K., R.G., N.J., E.K., N.P., N.N. and W.K.; visualization, S.H., N.K.K., P.M., W.D., N.K., R.G. and N.J.; supervision, W.K. and N.N.; project administration, W.K. and S.H.; funding acquisition, W.K. and S.H. All authors have read and agreed to the published version of the manuscript.

## Institutional Review Board Statement

This study was classified as exempt research by the Faculty of Associated Medical Sciences Ethic Committee (AMSEC-64EM-028).

## Informed Consent Statement

Not applicable.

## Data Availability Statement

Not applicable.

## Acknowledgments

We thank Dr. Barry Lutz and his laboratory members at the Department of Bioengineering at the University of Washington for their prior contributions related to the RT-LAMP assay development. We thank the AMS-CSC for providing the samples and the Research Institute for Health Sciences, CMU for the laboratory facilities.

